# Policy & Organizational factors that Affect the Utilization of Health Services for Alzheimer’s Disease Among the Latino Community – The Primary Care Provider Perspective

**DOI:** 10.1101/2025.03.27.25324793

**Authors:** Maria Mora Pinzon, Diana Martinez Garcia, Jaime Perales-Puchalt

**Author notes:** **Corresponding author:** Jaime Perales-Puchalt, University of Kansas Alzheimer’s Disease Center, Fairway, KS 66205, USA. **Author Contributions: Maria Mora Pinzon:** Methodology, Formal analysis, data curation, writing – Original Draft, Writing – Review & editing. **Diana Martinez Garcia:** Methodology, visualization, Data Curation, Formal analysis, Writing – Review & editing. **Jaime Perales Puchalt:** Conceptualization, Methodology, Resources, Data collection, Validation, Writing – Review & editing, Supervision, Project administration, Funding acquisition. **Ethical Approval and Informed Consent Statements:** The University of Kansas Medical Center Institutional Review Board approved this project (STUDY00145615) on 4/20/2020. A Data User Agreement between the University of Wisconsin-Madison and the University of Kansas Medical Center was executed to allow the sharing of de-identified data for this analysis. The UW-Madison Health Sciences Institutional Review Board (IRB) determined that this research met the criteria for exemption. **Data availability statement:** Data is available upon reasonable request to the corresponding author at. **Disclosure of Use of AI:** During the preparation of this work, the author(s) used Grammarly and Microsoft Copilot to improve the manuscript’s readability and format. After using these tools, the author(s) reviewed and edited the content as needed and take(s) full responsibility for the publication’s content.

## Abstract

**Background:** Latino individuals bear a disproportionate burden of Alzheimer’s disease and related dementias (ADRD), with higher risk, underdiagnosis, and limited access to quality care. Primary care providers (PCPs) are crucial for early detection and management. However, organizational and policy factors significantly impact their ability to provide culturally competent and equitable ADRD care for this community. This study explores PCP perspectives on these organizational and policy factors to inform the development of accessible models that improve early diagnosis, preventive care, and quality of life for Latino individuals with ADRD.

**Methods:** We used thematic analysis to analyze qualitative interviews with 23 diverse PCPs across the USA. We recruited our sample using snowball sampling. We strengthened the validity of our findings by using rigorous data reduction techniques.

**Results:** Key themes emerged highlighting the interplay of organizational and policy factors: 1) Insurance eligibility and care options for those uninsured were foremost, with mandated language services facing access and quality challenges that affected the ability of clinicians to perform an accurate diagnosis. 2) Staffing and available resources dictated the type of care offered, leading to inconsistent protocols and options. Providers reported that workup was influenced by their level of training, time availability, and comfort. 3) While recognized as crucial, comprehensive assessments that include evaluation of their home and social environment were limited by appointment constraints and lack of follow-up resources.

**Conclusion:** Economic and organizational factors, including insurance, costs, staffing models, and resource navigation, shape PCPs’ ability to deliver culturally competent and equitable ADRD care. Future interventions should address these barriers by training PCPs in diagnostic procedures in Latino communities and developing accessible service models and culturally appropriate diagnostic tools.

## INTRODUCTION

Currently, there is a deficiency in the workforce trained to treat dementia, including geriatricians,^1,2^ neurologists, psychiatrists, and other experts.^3-6^. It is estimated that in 2022, only 8,220 geriatricians were serving 52.4 million individuals over 65 years old, resulting in millions with limited access to a specialized diagnosis and management of cognitive symptoms. This has resulted in an increased burden to primary care providers (PCPs), with 82% of them being on the frontline of dementia care,^7^ but 40% reporting being uncomfortable making a diagnosis.^8^ Nearly half of PCPs sometimes choose not to assess a patient for cognitive impairment due to a lack of confidence in making a diagnosis and uncertainty around available resources,^9^ resulting in approximately 40% of older adults with probable dementia being undiagnosed.^10,11^

Despite PCPs’ pivotal role in the early detection and management of ADRD, there is evidence that various organizational and policy factors influence their ability to deliver culturally competent and equitable care. Understanding the extent of these factors within each community allows us to increase access to care. This study aims to fill this gap by exploring PCP perspectives on the organizational and policy barriers they face. By identifying these challenges, we can inform the development of accessible models that enhance early diagnosis, preventive care, and overall quality of life for Latino individuals with ADRD.

## METHODS

### Participant Recruitment and Data Collection

We have described the methods of the current study in detail elsewhere.^12^ To summarize, This is an analysis of a larger studies that conducted qualitative interviews to PCPs about ADRD care in the Latino community. PCPs were recruited using snowball sampling techniques. Inclusion criteria included being a physician (MD or DO), nurse practitioner, or physician assistant who currently or recently provided primary care services to Latino families with Alzheimer’s disease and related dementias (ADRD) in the U.S.

### Theoretical Framework

Andersen’s Behavioral Model of Health Services Use^13,14^ establishes that health outcomes result from contextual and individual factors that interact at the community, organization, and individual levels. Contextual characteristics relate to the circumstances and environment of healthcare access and include community and healthcare organization characteristics. Individual characteristics include genetic factors, social support structures, economic factors, and beliefs. Andersen’s model considers organizational factors either as contextual or individual characteristics. Contextual organizational factors refer to the amount, distribution, and processes for delivering services in a community, including outreach programs and office locations. Individual organizational factors refer to whether an individual has a primary care provider and how they interact with that provider (e.g., waiting time for care). Although Andersen’s model has been widely used to explain access to care, definitions for its domains vary across studies.^15^

### Data Analysis

Two reviewers independently conducted deductive qualitative content analysis. The Rigorous and Accelerated Data Reduction (RADaR) technique was employed to organize and code data in Excel.^16^ A research assistant performed initial open coding of the first five transcripts and reviewed them by an author to ensure accuracy. The themes were identified and refined through iterative coding and discussions. Consistency was maintained by dual coding every fifth transcript. A qualitative methodologist at the University of Wisconsin-Madison reviewed the final themes and codes.

### Ethics/Institutional Review Board Review

The University of Kansas Medical Center Institutional Review Board approved the project (STUDY00145615). Participants provided informed written consent online. A Data User Agreement facilitated the sharing of de-identified data between institutions. The UW-Madison Health Sciences Institutional Review Board determined that the research met exemption criteria.

## RESULTS

The analysis of qualitative interviews with 23 PCPs revealed five key themes **(Figure 1)**, organized across three categories in Andersen’s Behavioral Model of Health Services Use.^13,14^

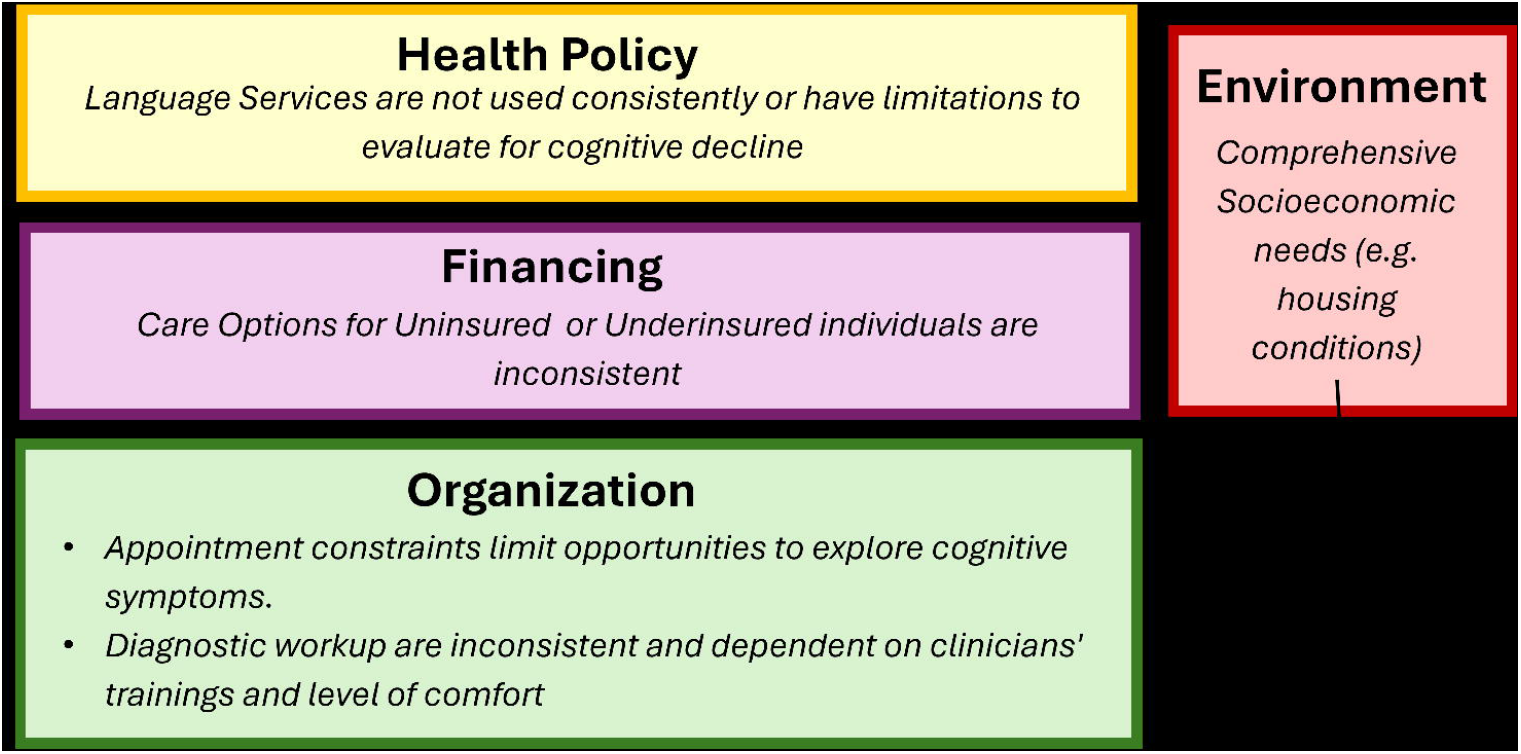

### Health Policy & Financing

PCPs reflected that mandated language services, while intended to improve care, often faced access and quality issues, hindering accurate diagnosis and effective communication with patients. PCPs also identified significant challenges related to insurance eligibility and care options for uninsured individuals, which required further assessment by people some team members. These constraints limit the ability of PCPs to provide comprehensive and culturally competent care (Table 1).

**Table 1:** Health Policy & Financing Elements That Affect Access To Healthcare Services According To Primary Care Providers.

| Theme | Quote |
| --- | --- |
| Language Services are not used consistently or have limitations to evaluate for cognitive decline | • "Yes, [language services] are free, there's no charge to the patient. The one problem that we have is that... the connection with the telephone sometimes is a problem. The nurses have to use their own cellphone for that, and I understand that is a problem but, even when they have the clinic cellphone, they are not friendly for that." (PCP 4)<br>• "Sometimes I usually try to use the Spanish that I have because sometimes it makes things easier and sometimes, I have noticed interpreters making errors, so it is just hard, it is not easy." (PCP 22)<br>• "I mean I do not really blame the provider because it takes a lot of time and these hospitals want you to see patients, so it like such a churn out that it takes a lot of time to get a translator, so some people just do the best they can without one unfortunately." (PCP 22) |
| Care Options for Uninsured or Underinsured individuals are inconsistent | • "[Donated specialty care program] is totally contingent upon how many providers are actually signing up for, how many patients they will take... It can take very long... [I] have had cases that take up to two years to even get an appointment and for neurology and neuropsychology it might be like six months." (PCP 12)<br>• "we have lots of strategies... It requires us to have a sense of what the family can do... she [care coordinator] qualifies every single patient, she looks at everyone's pay stubs, she computes the family household income... there's a lot that is sensitive private information, but as a physician it actually is very helpful, because... I don't have to know all the details, but at least if I have a sense of what they need, I can choose my therapy and it can fit their needs." (PCP 3) |

> *“It is over the phone, so it is less than optimal, to be honest. The phone is very hard, particularly with our population; many people have dementia and cognitive impairment and cannot hear, so it is very challenging. Hearing issues, cognitive issues, so the patient sometimes does not understand that it is a translator there and I am there like if it is on the phone you know with dementia it takes longer time processing*.*” (PCP 22)*

### Organizational Factors

The availability of staffing and resources dictates the type of care that can be offered, leading to inconsistent protocols and options across different practices. Many PCPs reported that their usual process includes ruling out reversible causes of cognitive decline such as anxiety, depression, medication interactions, and sleep apnea, followed by the use of cognitive screening tests such as the MOCA or SLUMS to decide if a referral to other services is needed. Meanwhile, other physicians commented that their ability to conduct thorough workups is influenced by their level of training, time availability, and comfort with ADRD care. Appointment constraints were a common theme across multiple providers, highlighting how expectations for 15–to 20-minute appointments limit their ability to investigate cognitive complaints in the presence of other complex conditions **(Table 2)**.

**Table 2:** Organizational Factors That Affect Access To Healthcare Services According To Primary Care Providers.

| Theme | Quote |
| --- | --- |
| Appointment constraints limit opportunities to explore cognitive symptoms. | • So, every visit is 15 minutes, when I have Spanish-speakers visit usually end up longer because they love talking to somebody who speaks their own language, so they always like to extend the visit. If I have a new patient, I try to time myself sometimes for 30 minutes. The new patients will come for a lot of other medical problems that 15-20 minutes, it's actually... it's really pushing it. (PCP 4)<br>• The average visit is 15 minutes... someone comes in with a list of six to seven issues and there is no way we can address that, but we try. I do primary care, some are quick and some are not so quick... some are complex that require a lot of additional management and lot of you know thinking and planning and time is always valuable. (PCP 8)<br>• They [Administration] would look at our numbers and be like you can do a new patient in 30 minutes, you can do an established patient in 15 minutes. My partner did do this, and I mean he is a great physician, but when I would see his patients for follow up, they would have no idea what he had said or what was recommended because he did not have time to address that. Even in a 30-minute visit if the patient has a lot going on it is just not enough time. (PCP 13) |
| Diagnostic workup are inconsistent and dependent on clinicians' trainings and level of comfort | • "I am the newest physician there and everybody does whatever they think is more appropriate for their own practice. So, nobody tells you what to do, and have to do things or how to report that. If somebody prefers to do this panel and run heavy metals, and everybody who is acting up, they do it, and if somebody feels that I could just tell you that you have dementia because you forgot your name today, then that's the diagnosis." (PCP 4)<br>• "We do not have a set protocol for anybody, we do not kind of training together, so we tend to bounce ideas off of each other, so I think our practices actually are pretty similar, but we do not have a defined protocol." (PCP 9)<br>• It depends on whether the patient is telling me to investigate. What we do try to address is depression and anxiety, basically what the USPSTF recommends (PCP 7).<br>• "I am much more equipped and comfortable with management of anything else, you know diabetes, hypertension, hyperlipidemia, you know those are fine. My geriatric population is small, but building, and I imagine as I age my patient base also ages and as they age and have more issues I also age and learn more about their issues." (PCP 8) |

### Need – Environmental factors

While PCPs recognized the importance of comprehensive assessments that included evaluations of patients’ home and social environments, these were often constrained by a lack of follow-up resources. This limitation hindered the ability to fully understand the multifaceted needs of Latino patients with ADRD, including their need for housing, assistance for activities of daily living, and safety. This limitation limited their capacity to provide resources for individuals **(Table 3)**.

**Table 3:** Environmental Factors That Affect Access To Healthcare Services According To Primary Care Providers.

| Theme | Quote |
| --- | --- |
| Comprehensive Socioeconomic needs (e.g. housing conditions) and connecting patients to local resources is a common part of the care plan | • "I wanted someone to come out to their house [patients] to make sure they were safe in their home... there was a few people I was really worried about, a whole discussion on family and there was also a lot of cohabitation and unhealthy living situations in the small towns I was in, there were some people where they were taking advantage of their elders." (PCP 13)<br>• "The thing that occurs with dementia ends up just being who is going to cook and clean, who keeps them independent as best we can, and activities of daily life." (PCP 8)<br>• "There is a lot of pain physical and I cannot say the top one, but probably functional because as dementia worsens of course you know the functional status worsens and the way our society is, is that a lot of times people live alone, well, Latino people actually often live with their families, which is good, so home attendant is very important or some sort of caregiver is very important, that is like probably the number one." PCP 22<br>• "[Need] resources for them to have at home, resource for you know pretty much all the tools they need to be safe in their own home or to be able to you know have that discussion of like whether you should go to a facility or be at home, yeah it would be nice to be in contact with you know social services or whatever the community has." (PCP 13)<br>• "I know that there was some like support groups specifically in the [City] place and then also there were companies that would actually come to the home and help, so I think that types of people would come and like help them do their activities of daily living and things around the house or maybe even take them to the grocery store or things like that." (PCP 17) |

> *“I don’t have anything. I don’t have anything that someone can take away with them and read or read again later with other family members. There’s nothing tangible I can give anybody right now*.*” (PCP 3)*

## DISCUSSION

This study highlights the significant impact of organizational and policy factors on the provision of ADRD care for the Latino community. This is one of the few studies in the Latino community that focuses on exploring the role of organizational and policy factors in accessing healthcare, as most studies take a rather individual approach.^17-19^ The findings reveal key themes related to policy constraints, resource-dependent care, comprehensive assessments, and family involvement, which collectively shape the ability of PCPs to deliver culturally competent and equitable care.

The study found that insurance eligibility poses substantial barriers to effective ADRD care. Previous research shows that PCPs treating Medicaid-insured and uninsured patients face resource constraints, limiting access to specialized diagnostics or multidisciplinary care coordination due to administrative barriers.^20^ Medicare coverage enables basic diagnostic imaging and specialist referrals, but PCPs report challenges in navigating prior authorization requirements and coverage limitations for newer therapies or biomarker testing, which delays diagnosis and complicates treatment plans.^20,21^ For uninsured or underinsured patients, PCPs often prioritize cost-sensitive approaches, such as deferring advanced neuroimaging or relying on non-pharmacological interventions.^22^

Furthermore, regarding language services, the Civil Rights Act of 1964 and Executive Order 13166 require healthcare organizations that receive federal funds, such as through Medicare or Medicaid/CHIP to provide interpreter services for Limited English Proficiency (LEP) individuals. Despite this legal requirement, it is known that not all hospitals comply; in 2010, 13% of hospitals fully complied with all National Standards for Culturally and Linguistically Appropriate Services (CLAS).^23^ Our study also reflects that for cognitively impaired patients, the risk of miscommunications due to inconsistent use of interpreters might increase the risk of errors in medication instructions or diagnostic clarity, which can directly harm individuals with diminished capacity to self-advocate or correct misunderstandings.^24^

The availability of staffing and resources dictated the care offered, leading to inconsistent protocols and options. This is consistent with studies that highlight the challenges faced by caregivers in resource-limited settings.^25,26^ Integrating emerging diagnostic biomarkers and comprehensive care paradigms can enhance the evaluation and management of ADRD in primary care settings.^27^ PCPs recognized the importance of comprehensive assessments, including evaluations of patients’ home and social environments. However, appointment constraints and lack of follow-up resources limit their ability to conduct thorough assessments. This finding is supported by research emphasizing the need for person-centered assessments incorporating modifiable and non-modifiable risk factors. Structured interventions that support family caregivers and enhance their engagement can significantly improve the quality of ADRD care, but accessing this is also limited by financing options. Insurance type also affects care continuity: fee-for-service Medicare patients with dementia experience higher hospitalization rates compared to those in managed care plans, underscoring the role of insurance structures in care stability.^28^

### Limitations of the Study

This study has several limitations, including the sample size and geographic scope, which may affect the generalizability of the findings. Although the study participants were recruited from various regions across the USA, the geographic distribution may not fully capture regional differences in healthcare delivery and policy environments. Furthermore, healthcare policies and organizational practices are dynamic and can change over time. Additionally, our study does not include the views of other stakeholders, such as patients, family members, or policymakers, and using snowball sampling techniques, while effective for recruiting participants, may introduce selection bias, as PCPs who chose to participate might have specific characteristics or experiences that differ from those who did not.

### Implications for future research and public health impact

The current study has implications for future research. We have identified five themes across three categories in Andersen’s Behavioral Model of Health Services Use. Future studies should assess the prevalence of these themes and their impact on the health of Latino families with ADRD. While cross-sectional studies are ideal to assess prevalence, longitudinal studies that track changes in PCP practices and policy impacts over time would provide a more comprehensive view of the evolving landscape of ADRD care in this community. PCPs highlighted the need for resources for Latino families with ADRD. While culturally tailored evidence-based interventions exist,^29^ these might not be ready to implement in the real world. Future studies should explore how to best implement these evidence-based interventions in primary care. Our study also has implications for public health. Issues with language services highlight the importance of integrating these services in a way that they do not impact the workflow. In-person and videocall options are preferred to phone call options. Our findings also suggest that it will be important to standardize assessments so that all, and not just those with optimal healthcare coverage have access to the gold standard. Minimum standards for ADRD care also need to be developed and disseminated to ensure that all primary care clinics, irrespective of their budgets, have the capacity to provide appropriate ADRD care to Latino families. These standards might have to include available remote options and workups that low-resource clinics can conduct or easily refer to. The National Institute of Health and the Alzheimer’s Association have free materials for Latino families, some of which have shown to reduce depressive symptoms among caregivers.^29,30^ All clinics could, in theory, request these to hand them out to their Latino patients and their families.

### Conclusion

Addressing organizational and policy barriers is essential for improving ADRD care for the Latino community. Targeted interventions that enhance PCP training, develop accessible service models, and promote family involvement can significantly improve early diagnosis, preventive care, and quality of life for individuals with ADRD. Further studies are needed to explore the cost-effectiveness of new policies and protocols that address the identified barriers and improve care delivery.

## Data Availability

Data is available upon reasonable request to the corresponding author at.

## Acknowledgments

Dr. Perales-Puchalt thanks the national and local organizations that have partnered with him to conduct present and past research since 2015. The research team thanks research participants included in all stages of this research as well as anyone who has contributed directly and indirectly to this research. The ideas and opinions expressed herein are those of the authors alone, and endorsement by the authors’ institutions or the funding agency is not intended and should not be inferred.

